# Exploring RSV transmission patterns in different age groups in the United States

**DOI:** 10.1101/2024.12.23.24319532

**Authors:** Ke Li, Virginia E. Pitzer, Daniel M. Weinberger

## Abstract

Respiratory syncytial virus (RSV) infections are a significant public health concern for pediatric populations and older adults, with seasonal winter outbreaks in the United States (US). Little is known about the timing of RSV epidemics across age groups and the relative contribution of within-group and between-group transmission of RSV in each age group. The lack of understanding of age-specific RSV transmission patterns limits our ability to inform vaccination policies. In this study, we examine the timing and transmission patterns of RSV epidemics across different age groups in 12 US states from 2018 to 2024. We found that infants under 1 year and young children aged 1–4 years experienced the earliest epidemic timing, while the elderly group had the latest. Using a semi-mechanistic age-structured spatiotemporal model, we further showed that between-group transmission greatly contributes (>50%) to the burden of RSV hospitalizations for children under 1, school-age children aged 5-17, and adults aged 18-64. By contrast, incidence in the elderly group (above 65 years) was primarily driven by transmission within the age group. Our findings indicate that distinct age groups play unique roles in propagating RSV epidemics in the US, with age-specific transmission patterns that can guide more effective RSV vaccination policies.

## Introduction

Respiratory syncytial virus (RSV) infections pose a major public health threat, especially for young children, older adults, and immunocompromised individuals [1,2]. An estimated 100,000 deaths of children under the age of 5 were attributed to RSV globally in 2019 [3]. In the United States (US), seasonal RSV epidemics exhibit consistent onset and peak timing, with temporal variations in different regions [4,5]. The epidemics typically begin in the Southeast, occur later in the Northeast and Midwest, and eventually end in the Southwest/West regions. While children aged 3–6 years are believed to play a prominent role in propagating RSV epidemics [6], little is known about the timing of RSV epidemics in each age group. The relative contributions of within-group and between-group transmission to RSV epidemics in each age group have also received limited attention.

There are currently a variety of RSV prevention products available, including vaccines for older adults and long-lasting monoclonal antibodies and maternal vaccination to protect young infants [7]. Anticipating age-specific timing and transmission patterns of RSV epidemics is important for informing how to most effectively use these new products and reduce the burden on high-risk populations. Mathematical models and statistical methods [4,5,8–10] have been used to study RSV transmission dynamics, evaluate the impact of interventions, and predict epidemic patterns, providing valuable insights into the mechanisms driving RSV seasonality and the potential outcomes of vaccination or other public health strategies.

In this work, we calculated the peak timing of RSV epidemics across different age groups across 12 states in the US between 2014 and 2018. We then applied a semi-mechanistic age-structured spatiotemporal model [11] to estimate the extent to which disease incidence in each age group can be linked to previous cases within the same group and across other groups, adjusting for the population distribution and reporting fraction for each age group. Finally, we discuss the relevance of our findings for informing vaccine administration.

## Methods

### Hospitalization Data

Weekly laboratory-confirmed RSV-associated hospitalization rates (per 100,000) for different age groups between 2018-2024 were obtained from The Respiratory Syncytial Virus Hospitalization Surveillance Network (RSV-NET). The data are drawn from 161 counties and county equivalents in 13 states participating in the Respiratory Virus Surveillance Network and can be obtained from https://www.cdc.gov/rsv/php/surveillance/rsv-net.html. The formatted data can be downloaded from https://github.com/keli5734/RSV_timing_agegroup. An RSV season is defined as starting from July of one year to June of the following year. In our analysis, we excluded North Carolina because it only contained data for the 2024/25 season.

### Demographic Data

To calculate the number of residents in a surveillance area hospitalized with laboratory-confirmed RSV based on the hospitalization rates (per 100,000), we multiplied the rates by the total population estimate for the surveillance catchment area (see https://www.cdc.gov/rsv/php/surveillance/rsv-net.html for detailed catchment areas in each state). For calculations prior to the 2020–2021 season, bridged-race population estimates from the National Center for Health Statistics were used as multipliers. The data can be found in https://www.cdc.gov/nchs/nvss/bridged_race.htm. Beginning with the 2020–2021 season, unbridged census population estimates (U.S. Census Bureau, Population Division, Vintage 2020–2022 Special Tabulation) have been used instead.

### The center of gravity and the intensity of RSV activity

The timing of the seasonal epidemics can be summarized using the center of gravity. The center of gravity of RSV activity for each season (*G_s_*) was measured as the mean epidemic week, with each week weighted by the weekly number of hospitalizations (*H*_*s*,*w*_), such that 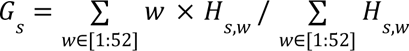, where *w* is an index for the week number in each epidemic season, *s*. *w* = 1 indicates the first full week of July in the previous year, and *w* = 52 indicates the last full week of June in the following year.

### The effective reproduction number

We used the *EpiEstim* package (version 2.2-4) [12] in R (version 4.3.2) to estimate the time-varying effective reproduction number during the seasonal epidemics. We first interpolated the weekly hospitalization data to estimate daily values, which were used as the inputs. We assumed the mean serial interval of RSV infection to be 7.5 days, with a standard deviation of 2.1 days, based on a review [13]. We set the starting day to the 60th day of the entire time series, corresponding to early September 2018, and used a moving window of 250 days to reduce estimation uncertainty.

### Mathematical model

To estimate the proportions of RSV hospitalizations attributable to within-group transmission and between-group transmission, we applied a framework from the *hhh4contacts* package (version 0.13.3) in R (version 4.3.2) [11]. This approach models the number of cases in an age group at a specific time as a function of the number of cases in a previous time point from the same age group or other age groups, as well as covariates for the rate of hospitalizations and the autoregressive components. The number of reported hospitalizations in a specific age group *g* at time *t*, *H*_*gt*_, is assumed to follow a negative binomial distribution with mean µ_*gt*_ and an age-specific overdispersion parameter Ψ_*g*_, such that *H*_*gt*_∼*NegBin*(µ_*gt*_, Ψ_*g*_). The mean µ_*gt*_ is additively decomposed into endemic (*e*_*gt*_ *v*_*gt*_) and observation-driven epidemic components 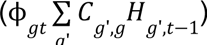, such as

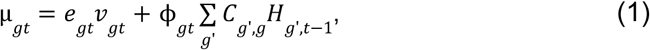

where *c*_*g*’,*g*_ is the contact matrix between age group *g*’ and *g*. The contact matrix was obtained from [14] where age-specific contact patterns were estimated in the US (see **SFig. 1**). The log-linear predictors for the endemic component are given as:

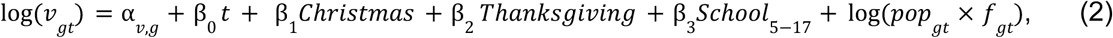

The predictors for the epidemic component are given as:

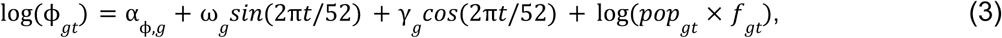

Where α_*v*,*g*_ and α_ϕ,*g*_ are age-specific intercepts for each component. *christmas*, *Thanksgiving* and *school*_5−17_ are dummy variables indicating the weeks of Christmas (the last week of December), Thanksgiving (the last week of November) and school holidays (based on public school calendars in different states, see https://github.com/keli5734/RSV_timing_agegroup). The terms *sin*(2π*t*/52) and *cos*(2π*t*/52) are age-specific seasonal forcing in the epidemic component. We also included age-specific population sizes (*pop*_*gt*_) and reporting fractions (*f*_*gt*_) as offsets in both the endemic and epidemic components. We fitted the model to each state individually. Note that we also tested and fitted different model structures to data, but the model shown above was the best based on Akaike information criterion (AIC) score.

### Model validation

To validate the *hhh4* model framework, we applied a simulation-and-estimation approach. We first fitted an RSV transmission model [5] to the hospitalization data for RSV in New York to estimate parameters related to the transmission rate, seasonal forcing and timing, duration of maternal immunity and reporting fractions using different contact matrices (i.e., a homogeneous contact matrix and a strongly assortative contact matrix). We simulated synthetic RSV data based on the best-fit model with the different contact matrices. We then used the model (**Eqs. 1-3**) to estimate the proportion of RSV cases resulting from within-group and between-group transmission based on the simulated data. We found that the proportion of cases due to within-group transmission increased for the data generated using the strongly assortative contact matrix, whereas the proportion of between-group transmission decreased. We also found that the best-fit model for the simulated data could only be obtained when applied to the corresponding contact matrix (see **SFig. 2**).

## Results

### RSV hospitalizations in different age groups

We stratified the total number of hospitalizations in each region into five age groups (**Fig. 1**). Across the five RSV seasons, children <5 years and older adults above 65 years accounted for the majority of hospitalizations in the Southeast (81.1%, **Fig. 1A**), Northeast (81.1%, **Fig. 1B**), Midwest (92.3%, **Fig. 1C**), and Southwest/West (83.1%, **Fig. 1D**). RSV activity was disrupted in the 2020/21 season due to the COVID-19 pandemic. In other seasons, RSV activity in the infant and young children group led that in other age groups, as indicated by early increases and peaks in RSV cases.

**Figure 1.**
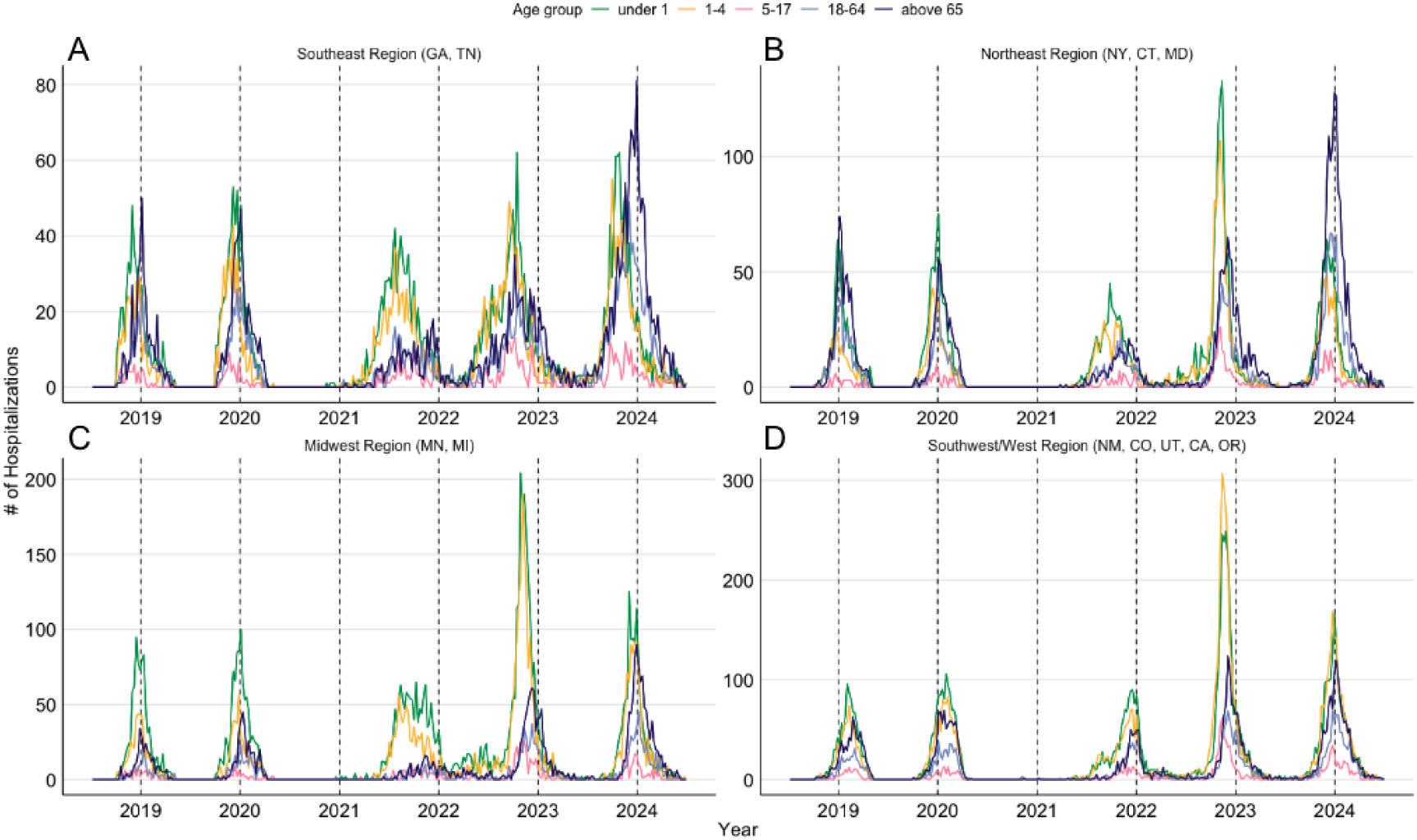
RSV-associated hospitalizations in different age groups. RSV hospitalizations were aggregated into (**A**) Southeast region (GA: Georgia, TN: Tennessee), (**B**) Northeast region (NY: New York, CT: Connecticut, MD: Maryland), (**C**) Midwest region (MN: Minnesota, MI: Michigan) and (**D**) Southwest/West region (NM: New Mexico, CO: Colorado, UT: Utah, CA: California, OR: Oregon), by state based on geographic location. Dashed lines indicate the first day of January of each year. The increase in hospitalizations between 2021 and 2024, particularly among older adults in the Southeast and Northeast regions, is likely due to the increased testing frequency in this age group. Hospitalization data at the state level is provided in **SFig. 3**.

### Timing of RSV epidemics differs among age groups

We next sought to quantify the timing of RSV epidemics in different age groups. To do this, we computed the center of gravity of each RSV season in each state. We observed that the timing is similar between the infant and young children groups, and this pattern remains across all seasons and states. This is evident as all points aligned along the diagonal line (**Fig. 2A**). All points shifted to the bottom-left corner due to an early out-of-season summer outbreak in the US during the 2021/22 season due to the COVID-19 pandemic.

**Figure 2.**
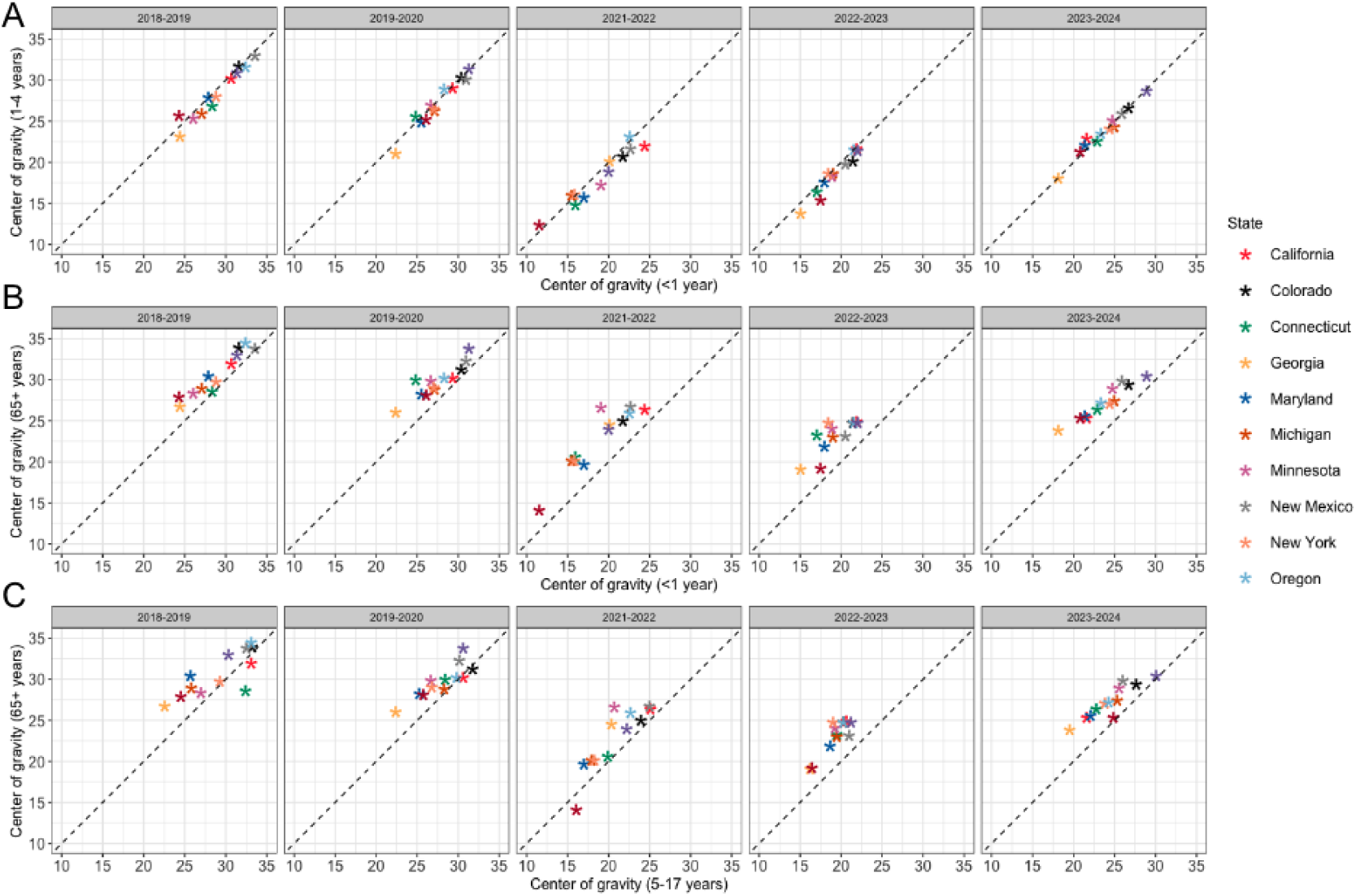
Timing of seasonal RSV activity in different states. Center of gravity of RSV activity—a measure of the mean epidemic week—was calculated for each season and state between **(A)** under 1 and 1-4 years, **(B)** under 1 and above 65 years, and **(C)** 5-17 years and above 65 years. Notice that the 2020-2021 RSV season was excluded due to the COVID-19 pandemic.

By contrast, the infant group has an early RSV epidemic compared to the elderly group in all states, with all points aligned above the diagonal line (**Fig. 2B**). Across all seasons and states, the epidemic in the infant group, on average, led the elderly group by 2.68 weeks. A similar pattern of RSV timing was also observed between the school-age children (5-17 years) and the elderly group (**Fig. 2C**). Two states, California and Connecticut, showed a later RSV epidemic in the school-age group compared to the elderly group during the 2018/19 season (as shown in the leftmost panel in **Fig. 2C**). This is because few cases were reported among school-age children in the later season (**SFig. 3**). In addition, we found a less consistent relationship of RSV timing between school-age children and young children across the states (**SFig. 4**). We observed that school-age children experienced relatively delayed RSV activity compared to infants and young children in the 2021/22 season, but not in other seasons. This could be explained by the quick rebound of RSV activity in the infants and young children after the COVID-19 pandemic due to increased susceptibility to RSV infections.

### The effective reproduction number of RSV epidemics

We also computed the age-specific effective reproduction number, *R*(*t*), for the seasonal RSV epidemics in each state to capture the temporal changes of RSV activity across age groups. If *R*(*t*) > 1, it indicates that RSV incidence is increasing; otherwise, the incidence is decreasing (i.e., *R*(*t*) < 1). We observed that although the estimated magnitude of *R*(*t*) is comparable among age groups, there is temporal variation between them (**Fig. 3** and **SFig. 5**). For the infants and young children groups, the effective reproduction number exceeds the threshold value (i.e., *R*(*t*) = 1) and reaches its peak earlier in each epidemic season, suggesting that these groups experienced early outbreaks in the beginning of each RSV season. In contrast, the older age group shows delayed increases in *R*(*t*), with peaks occurring later in the epidemic curve. The results demonstrate that transmission dynamics differ across age groups, with younger populations experiencing earlier initial spread, whereas the older age group shows later increases in RSV transmission.

**Figure 3.**
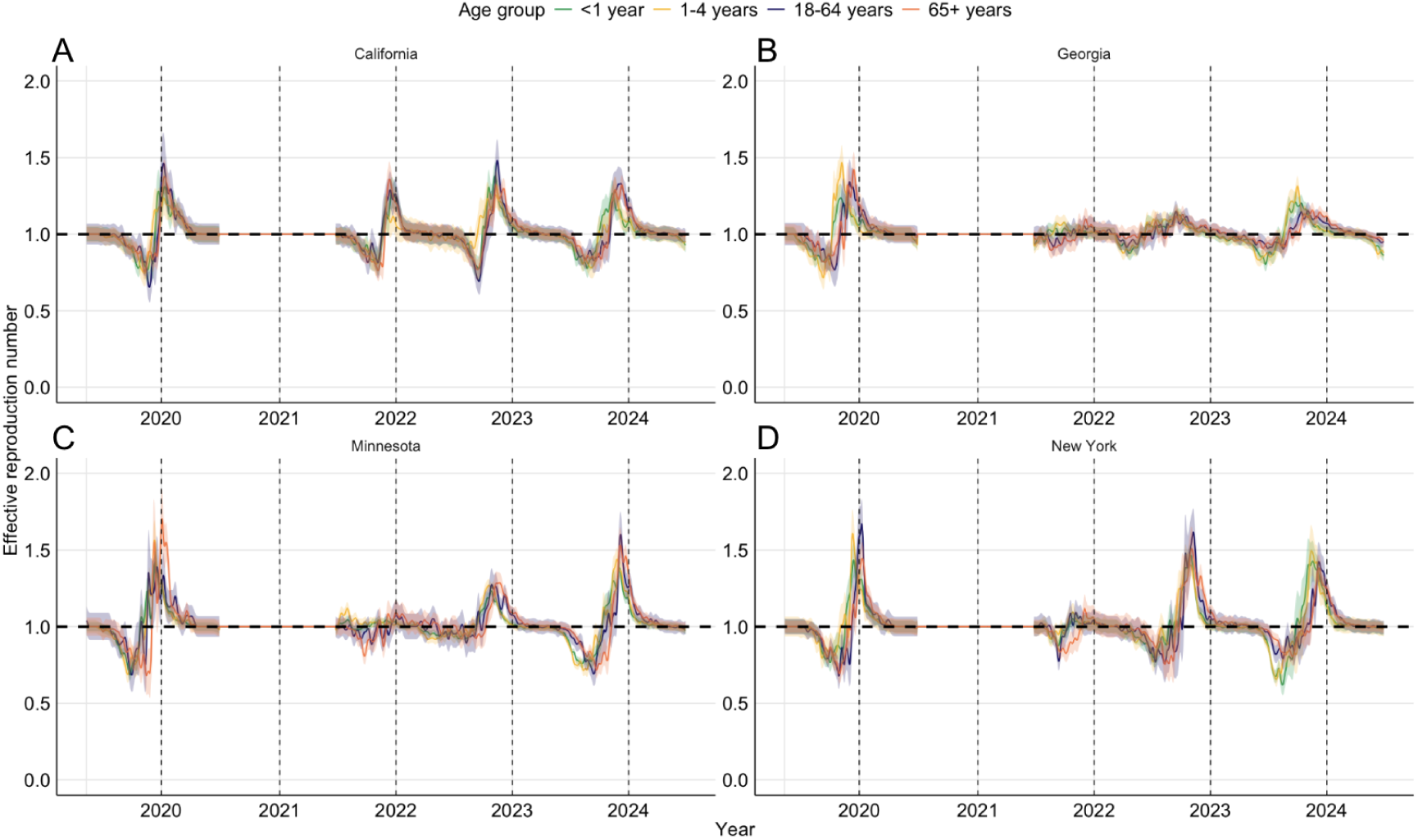
The effective reproduction number of seasonal RSV epidemics. **(A-D)** Solid lines indicate the mean estimates of *R*(*t*) for each age group, and the shaded area represents a 95% confidence interval for each age group. Dashed lines indicate the first day of January of each year. Note that *R*(*t*) was not estimated for the 2020/21 season, as no RSV hospitalizations were reported during this season due to the COVID-19 pandemic. Also note that *R*(*t*) was not estimated for the 5-19 age group, as few cases were reported. The estimated *R*(*t*) for other states is provided in **SFig. 5**.

### RSV hospitalizations due to within- and between-group transmission

Having demonstrated a leading RSV epidemic in the group aged under 5 years, we next estimated the proportions of RSV hospitalizations attributable to within-group transmission and to transmission from other age groups. We first fitted the model to hospitalization data from 15 counties in New York (**Fig. 4A**). The estimated hospitalization values allowed us to calculate the (time-averaged) proportions of the mean (i.e., µ_*gt*_) explained by the different components. We found that transmission within the same age group accounted for an estimated 32% and 42% of new cases in infants (**Fig. 4B**) and young children (**Fig. 4C**), respectively. By contrast, the proportion of within-group transmission was small among school-age children (**Fig. 4D**) and adults (**Fig. 4E**), accounting for only 14% and 21% of cases, respectively. In the elderly group, we found that the largest portion of hospitalizations, 68%, was attributed to transmission within the group (**Fig. 4F**). In addition, the endemic component contributed minimally to the observed hospitalizations across all age groups, accounting for less than 3%.

**Figure 4.**
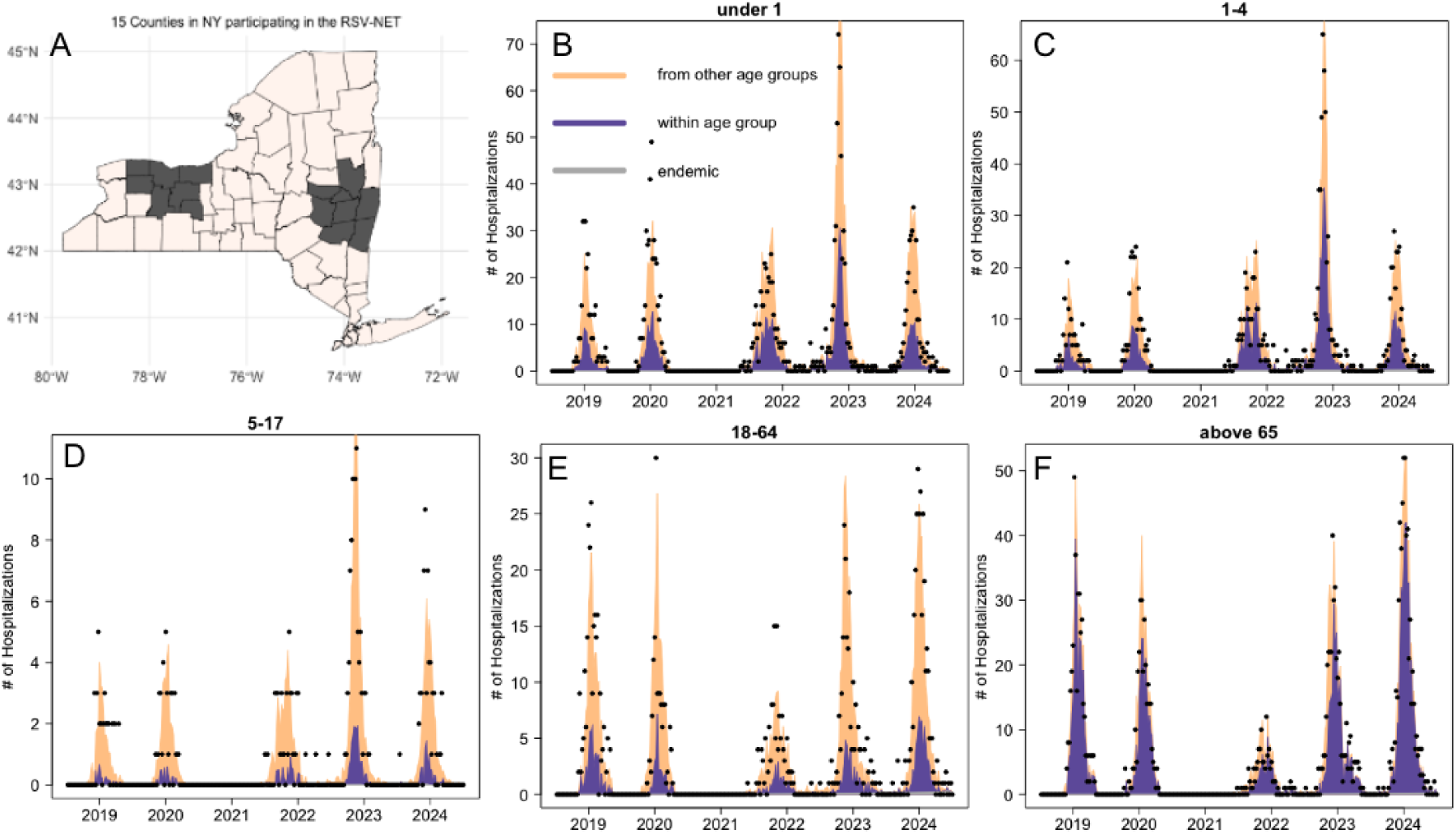
Age-specified RSV transmission pattern in New York. (**A**) Longitudinal hospitalization data were obtained from 15 counties in New York state participating in the Respiratory Virus Surveillance Network. (**B-F**) Fitted mean values of the epidemic component due to between-group (in orange) or within-group (in purple) transmission, and the endemic (in gray) component in the best-fit model based on AIC for the five age groups. Estimated parameter values are given in **Table S1**. Dots represent data points.

We also estimated the proportions of RSV transmission in other states. Across all states, we observed that transmission within the infant group accounted for an estimated 33.5% (95% confidence interval (CI): [24.9%, 42.1%]) of new cases in infants (**Fig. 5A**), while a larger proportion, 43.3% (95% CI: [28.5%, 58.1%]), was attributed to transmission within the group for young children aged 1–4 years (**Fig. 5B**). For school-age children and adults, we found that the majority of cases originated from other groups: 82.2% (95% CI: [76.9%, 87.6%]; **Fig. 5C**) and 79.4% (95% CI: [67.5%, 91.3%]; **Fig. 5D**), respectively. The proportions of cases due to within-group or between-group transmission in the elderly group varied across the 12 states, with a mean of 59.8% (95% CI: [35.5%, 84.2%]; **Fig. 5E**) attributable to within-group transmission.

**Figure 5.**
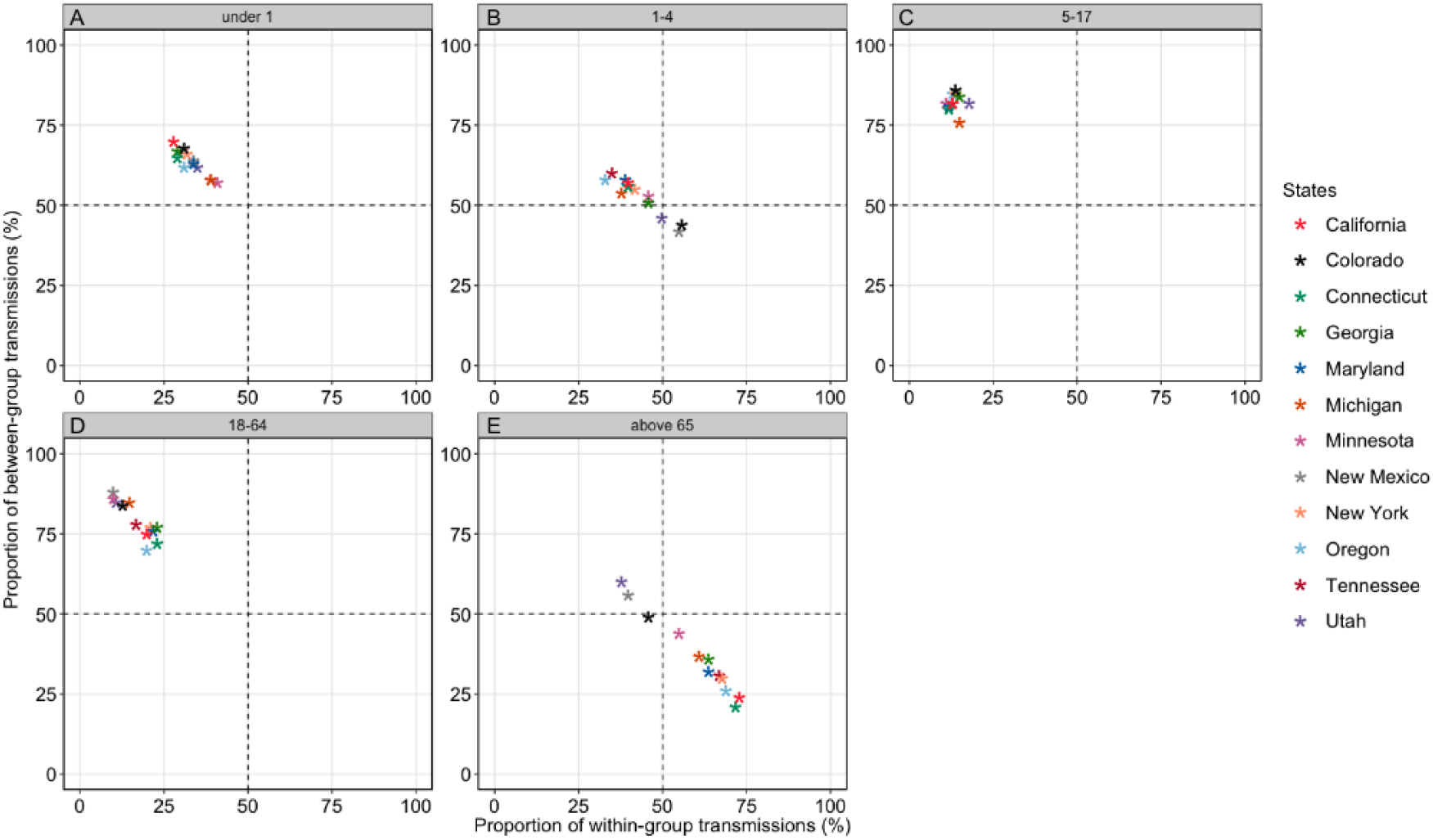
Proportions of within-group and between-group RSV transmission in different age groups. (**A-E**) The estimated hospitalization values from the best-fit model of each state were used to calculate the time-averaged proportions of the mean explained by within-host transmissions (horizontal axes) and between-group transmissions (vertical axes) in each age group.

There was some variation in the estimated proportions of within-group and between-group RSV transmission among young children (**Fig. 5B**) and the elderly (**Fig. 5E**) across the 12 states. We did not identify any covariates strongly associated with this variation. Neither state-specific daycare attendance rates nor average household sizes could explain the estimated differences. Contact patterns between age groups may vary across states; however, we could not test this due to the lack of state-specific contact matrices. Interestingly, we found a strong positive correlation between the proportion of within-group transmission in the 1-4 age group and the between-group transmission in those aged 65 and above (Pearson’s correlation coefficient *r* = 0.81, *p* < 0.005; **SFig. 6**).

## Discussion

In this work, we examined the timing of annual RSV epidemics in different age groups across 12 US states by calculating the center of gravity and the effective reproduction number for hospitalization data from 2018 to 2024. Although RSV transmission shows spatial and temporal variations across different regions in the US [4,5], the relative differences in epidemic timing between age groups remain consistent across states in all studied seasons, both before and following the COVID-19 pandemic. Our results suggest that children under 5 years old experience the earliest RSV epidemic, whereas the elderly group (those aged 65 and above) exhibits the latest onset and peak timing of RSV activity. In addition, we calculated the time-averaged proportion of RSV burden attributable to within-group or between-group transmission for each age group. Our model shows that hospitalizations among infants (under 1 year) and school children (5-17 years) were largely attributed to between-group transmission (>50%), while young children aged 1–4 years were more likely to acquire infections from their own age group compared to the other two groups. In contrast, the majority of cases in the older adult group were due to within-group transmission.

Previous work indicated that early RSV epidemics (in children under the age of 2) were associated with higher population density and larger household sizes in the area [9]. Here, we demonstrated that young children experience earlier timing compared to other age groups. The result does not necessarily indicate that young children introduce RSV into the population at the beginning of an epidemic. Rather, it suggests that during an observed epidemic, most cases occurred in the young age group, driving the early stages of the RSV epidemic. Although pinpointing the exact factors contributing to the early epidemic among young children is challenging, frequent contact within the group and their enhanced susceptibility to RSV infection may facilitate early viral spread and epidemic growth in the group. Future studies on RSV transmission that incorporate genomic analyses [15–18] could provide insights into the key drivers of transmission.

While different prevention strategies for severe infections in young children are currently available [19–22], the effectiveness of these measures in reducing disease transmission—either directly within their own age group or indirectly to other age groups—remains unclear. Understanding how RSV is transmitted within and between age groups can provide better insights into evaluating the effectiveness of these strategies. Our model results suggest that between-group transmission accounts for the majority of infections in infants. The results have important implications for planning vaccinations. Maternal vaccination for women during pregnancy and passive immunizations (e.g., monoclonal antibodies) for neonates and infants can protect the population by reducing transmission from other groups. Besides, passive immunization for eligible young children can provide direct protection to individuals in this age group, given a relatively high estimate of within-group transmission. Our results also show that within-group transmission drives the epidemic in the elderly group. This suggests that vaccination programs aimed at providing direct protection to the elderly population, thereby reducing the RSV burden within this group, may not be effective in significantly reducing cases in other age groups but could provide indirect protection to other older adults.

We found a positive correlation between the proportion of within-group transmission in the young children group and the proportion of between-group transmission in the elderly group across states. This suggests that states with higher transmission among young children also experience more of this transmission spilling over to the older age group. When RSV circulates heavily within the 1–4 age group, it likely increases the risk of transmission across age groups, particularly affecting individuals aged 65 and older and posing an increased risk for older adults. This pattern could reflect frequent interactions between the age groups, such as during family gatherings or in households where grandparents provide caregiving. The results also suggest that administering the RSV vaccine to these age groups could potentially help mitigate RSV epidemics in the elderly group.

There are some additional limitations to our study. Variations in testing practices across different states, seasons and age groups could have influenced the observed hospitalization rates, potentially introducing reporting biases that may not be fully accounted for in the model. Currently, our model only considers the time-varying, age-stratified reporting fractions of RSV infections for New York, which were previously estimated and assumed for all other states. In addition, our model assumes that the spatial spread of RSV in each age group follows a uniform diffusion process, which may not fully capture local variations in epidemiological factors, such as healthcare access and population differences or variations in susceptibility to infection across different age groups. Further studies that integrate more granular data could explore the impact of these factors on age-specific RSV transmission patterns.

Overall, our study sheds new light on the role of different age groups during RSV epidemics despite the above limitations. It reveals that children under the age of 5 years experience earlier RSV epidemics compared to other age groups, suggesting the prominent role of young children in driving seasonal RSV epidemics. We hope our study contributes to a better understanding and closer surveillance of the age-specific timing of seasonal RSV epidemics. Furthermore, estimates of age-specific RSV transmission patterns can be used to inform and evaluate the potential benefits of different RSV vaccination and prevention strategies for different population groups.

## Data Availability

All data produced are available online at https://github.com/keli5734/RSV_timing_agegroup

## Acknowledgments

This work was supported by a grant from the National Institutes of Health (R01AI137093). The content is solely the responsibility of the authors and does not necessarily represent the official views of the National Institutes of Health.

## Competing interest statement

DMW has received consulting fees from Pfizer, Merck, and GSK, unrelated to this manuscript, and has been PI on research grants from Pfizer and Merck to Yale, unrelated to this manuscript. The other authors declare no competing interests.

## Data and code availability

We used the R statistical software (v4.0.2) for all statistical analyses and visualization. Data and code used in this study are publicly available on Github: https://github.com/keli5734/RSV_timing_agegroup

## Supplementary Figures

**Supplement Figure 1.**
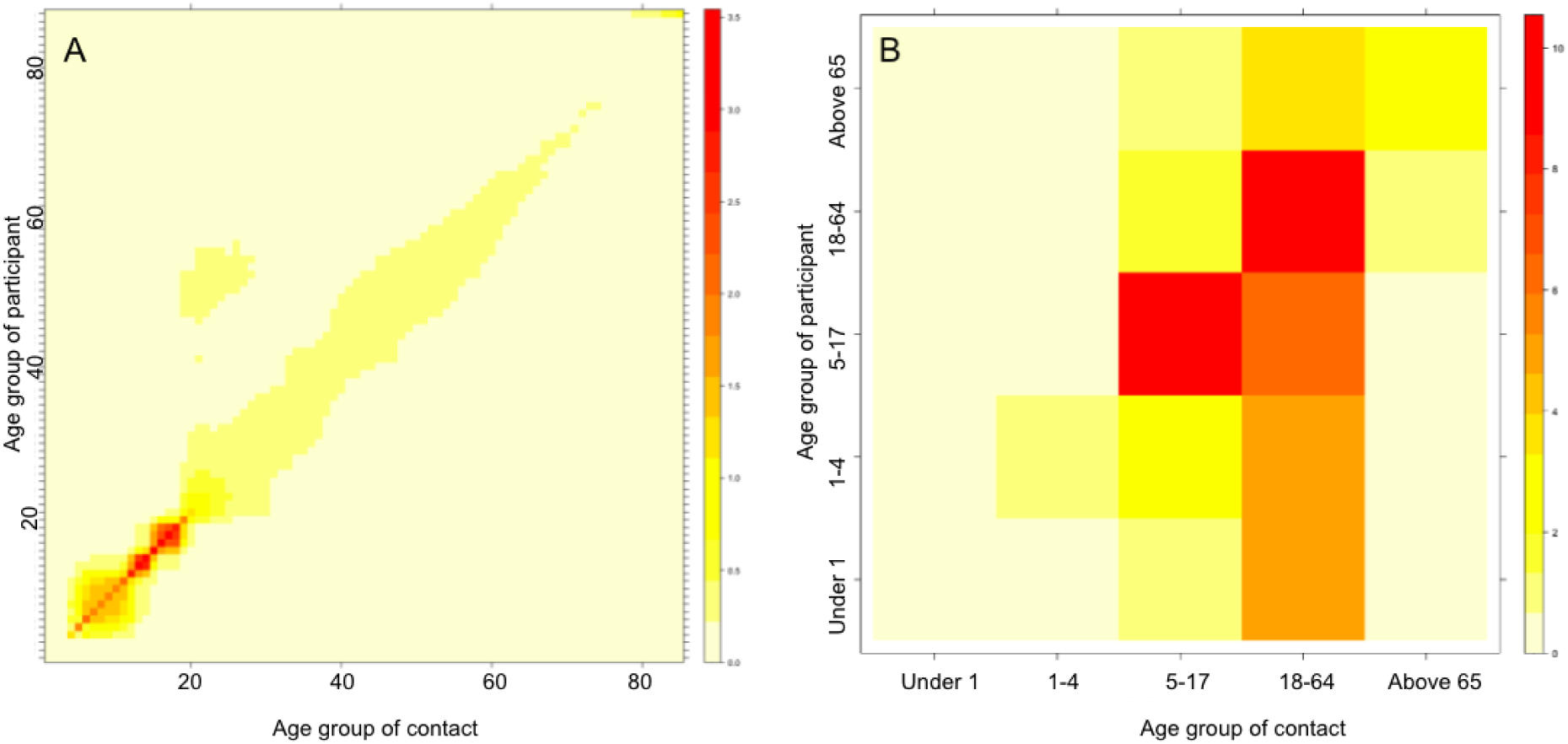
Human mixing patterns for disease modeling in the US. **(A)** Original contact matrix reflecting high-resolution mixing patterns (1-85 years) in the US. **(B)** Aggregated contact matrix reflecting mixing patterns between specified age groups in our study. Aggregation of the contact matrix is done by summing over the contact groups (columns) to be joined and calculating the weighted average across the corresponding participant group(rows), with weights equal to the group sizes.The aggregated contact matrix is asymmetric because of the different sizes of the involved age groups.

**Supplement Figure 2.**
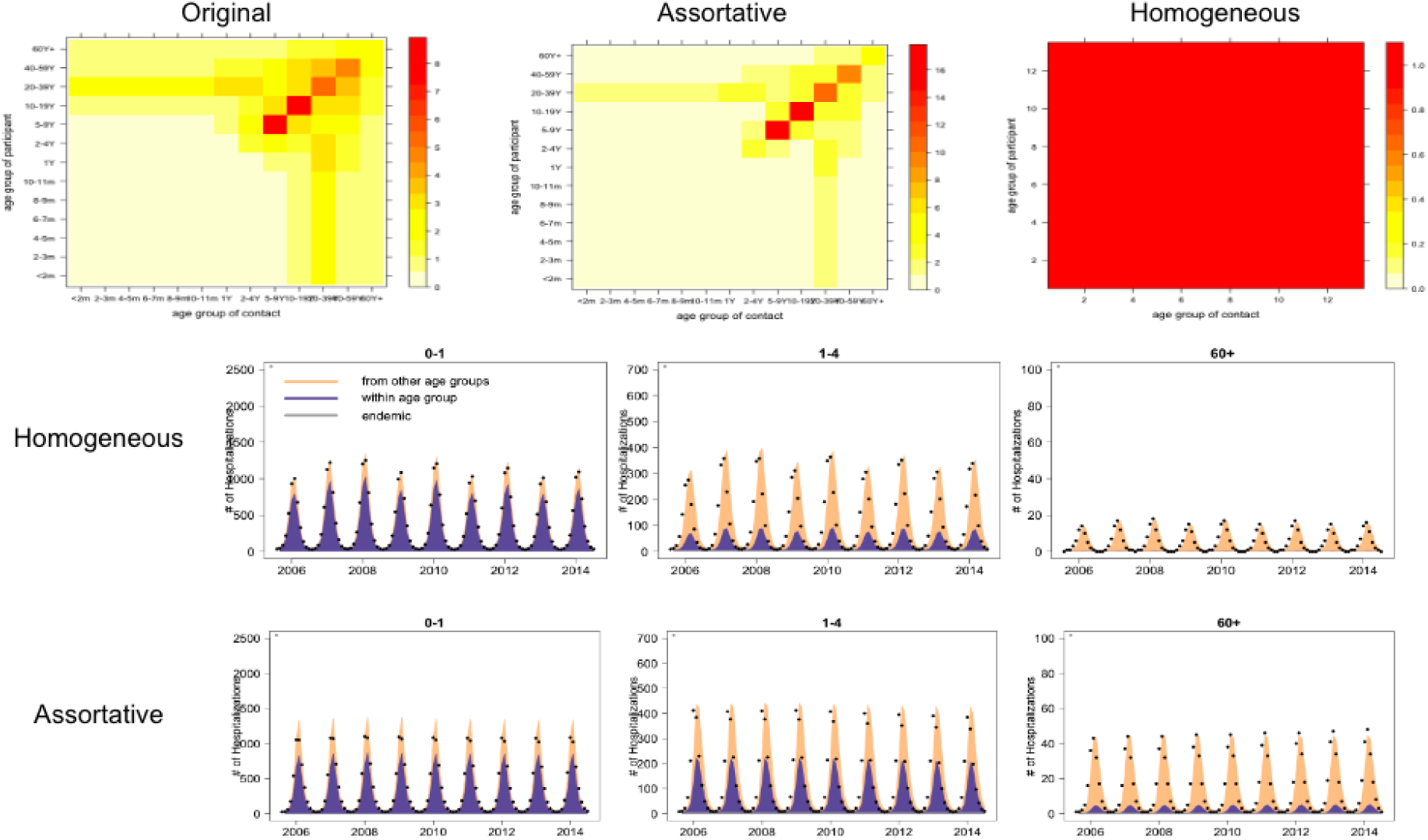
Model validation results. Fitted mean values of the epidemic component due to between-group (in orange) or within-group (in purple) transmission, and the endemic component (in gray) in the best-fit model based on AIC, using different contact matrices. The contact matrices reflect mixing patterns between 13 age groups. Dots represent simulated data points from an RSV transmission model with the corresponding contact matrices. The assortative and homogeneous matrices were derived based on the original contact matrix.

**Supplement Figure 3.**
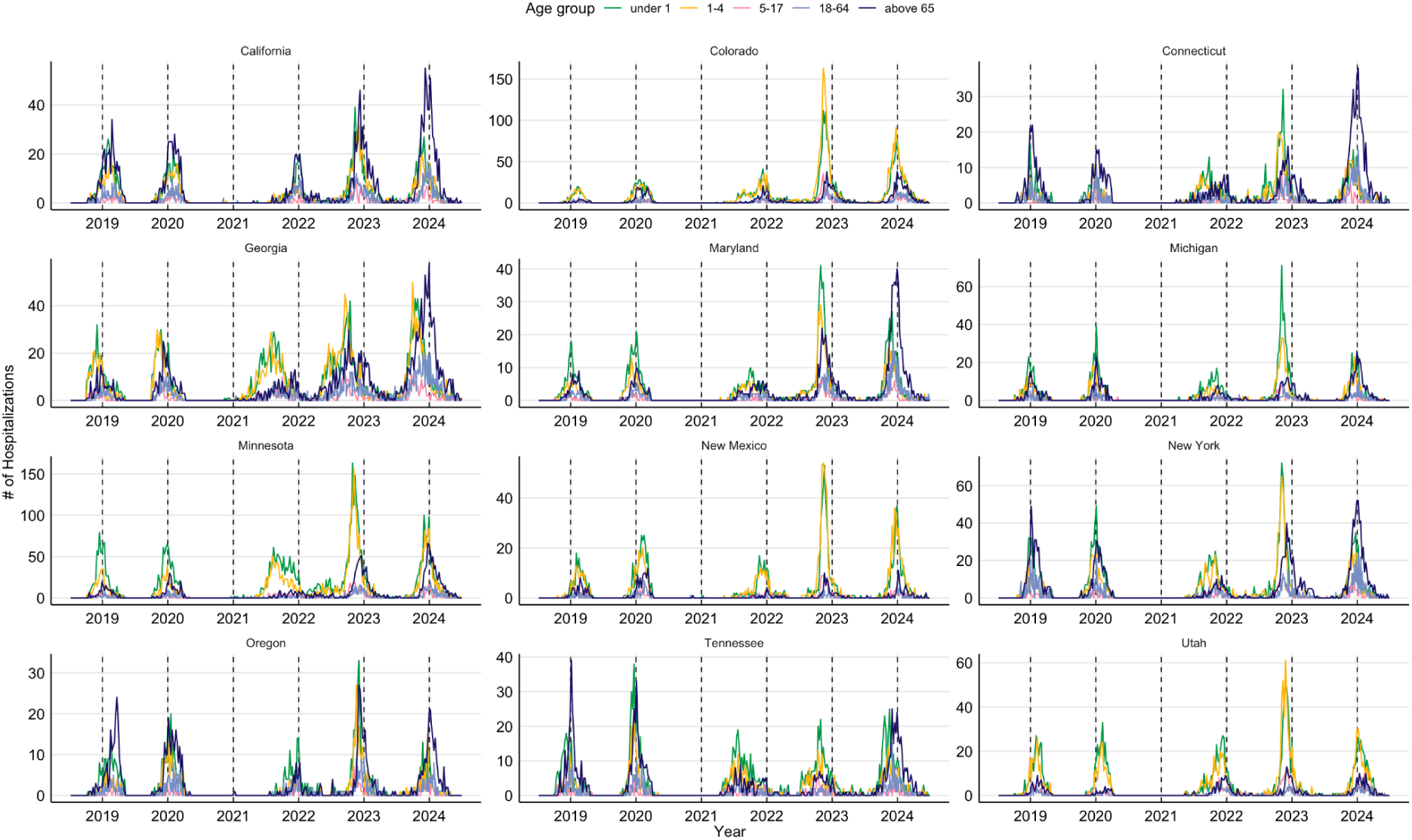
RSV-associated hospitalizations in different age groups. RSV hospitalizations from the 12 US states participating in the Respiratory Virus Surveillance Network. Dashed lines indicate the first day of January of each year.

**Supplement Figure 4.**
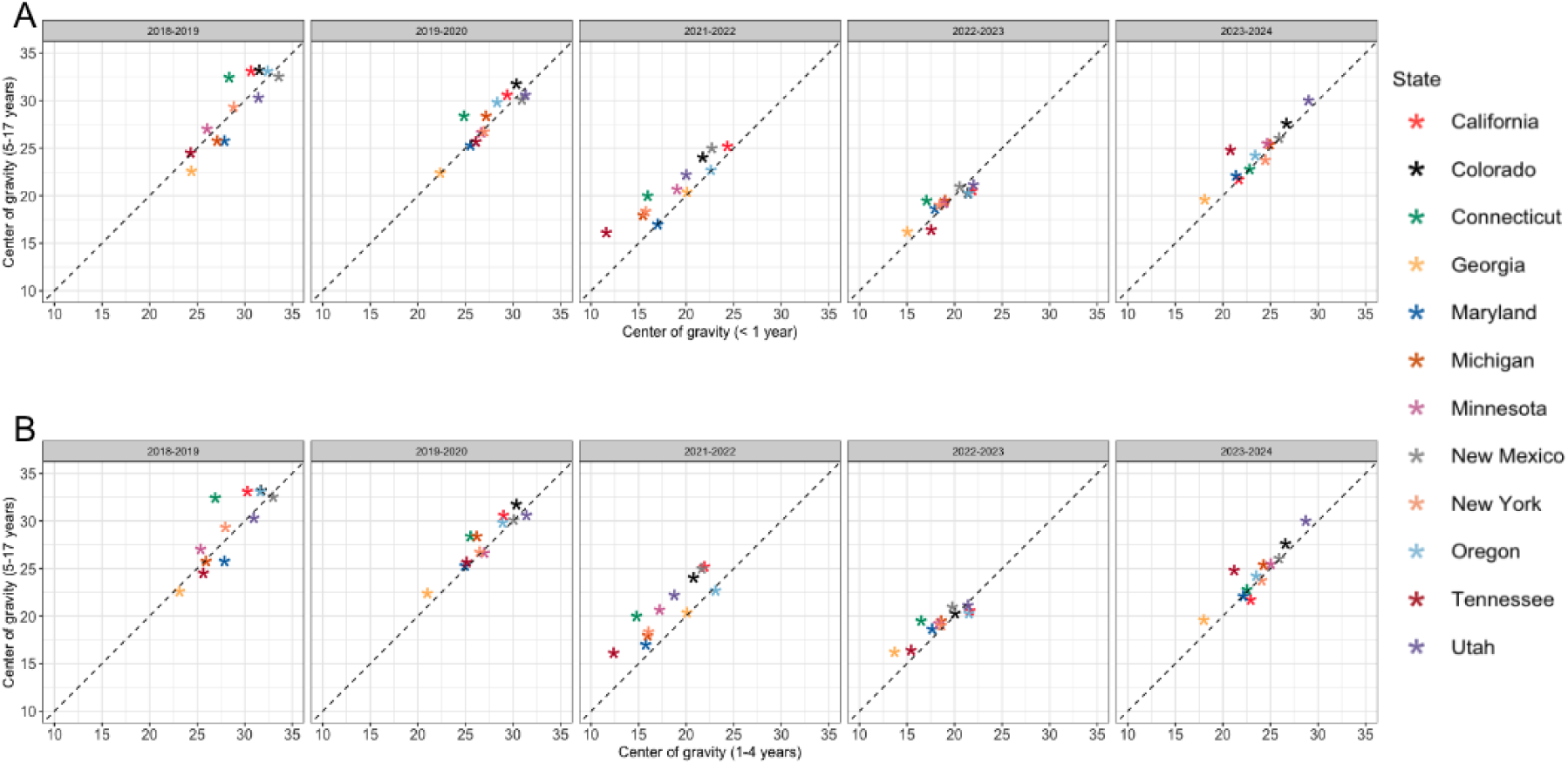
Timing of seasonal RSV activity in different states. Center of gravity of RSV activity—a measure of the mean epidemic week—was calculated for each season and state between **(A)** under 1 and 5-17 years, and **(B)** 1-4 and 5-17 years. Notice that the 2020-2021 RSV season was excluded due to the COVID-19 pandemic.

**Supplement Figure 5.**
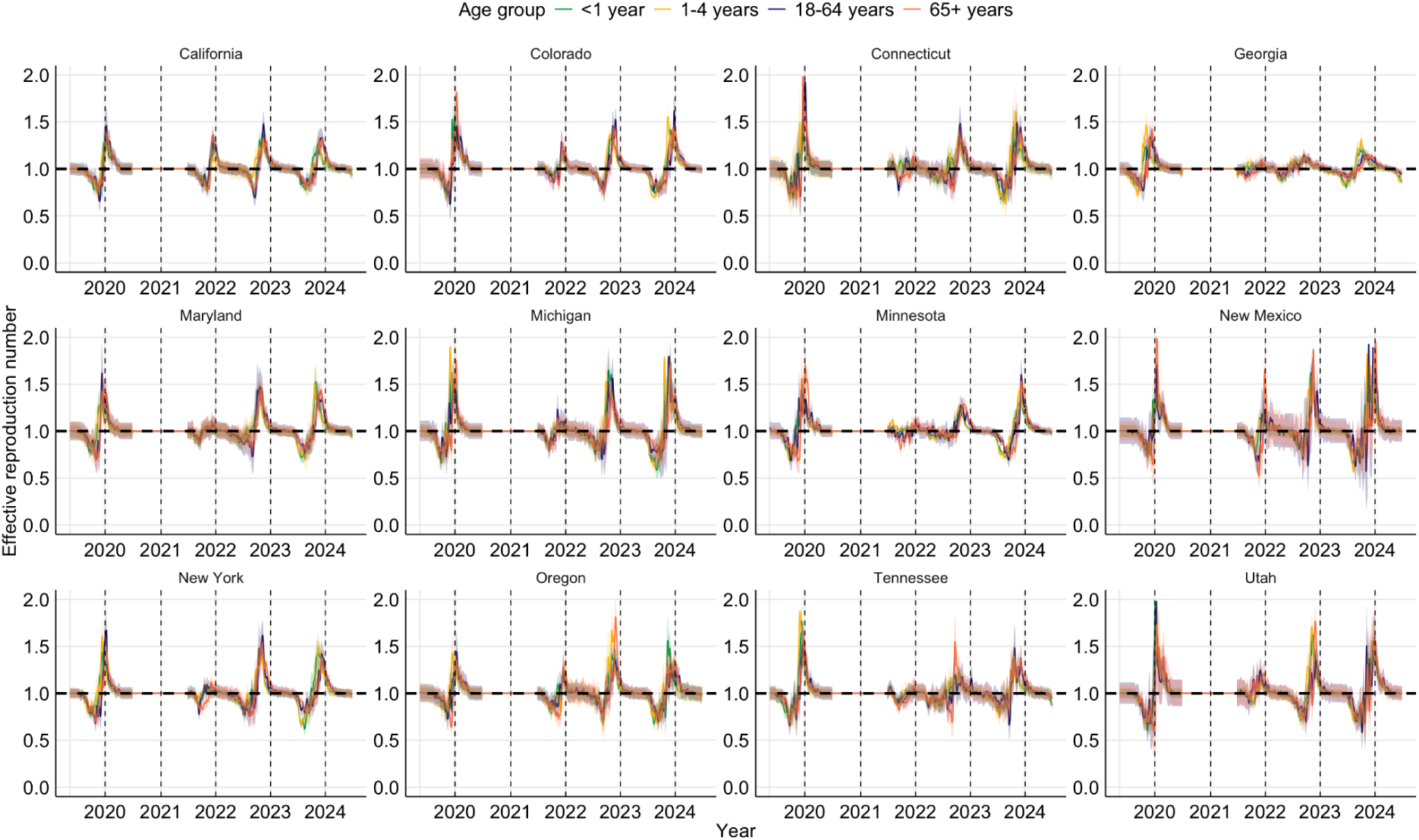
The effective reproduction number of seasonal RSV epidemics. Solid lines indicate the mean estimates of *R*(*t*) for each age group in 12 US states, and the shaded area represents a 95% confidence interval for each age group. Dashed lines indicate the first day of January of each year. Note that *R*(*t*) was not estimated for the 2020/21 season, as no RSV circulated during this season due to the COVID-19 pandemic. Also note that *R*(*t*) was not estimated for the 5-19 age group, as few cases were reported.

**Supplement Figure 6.**
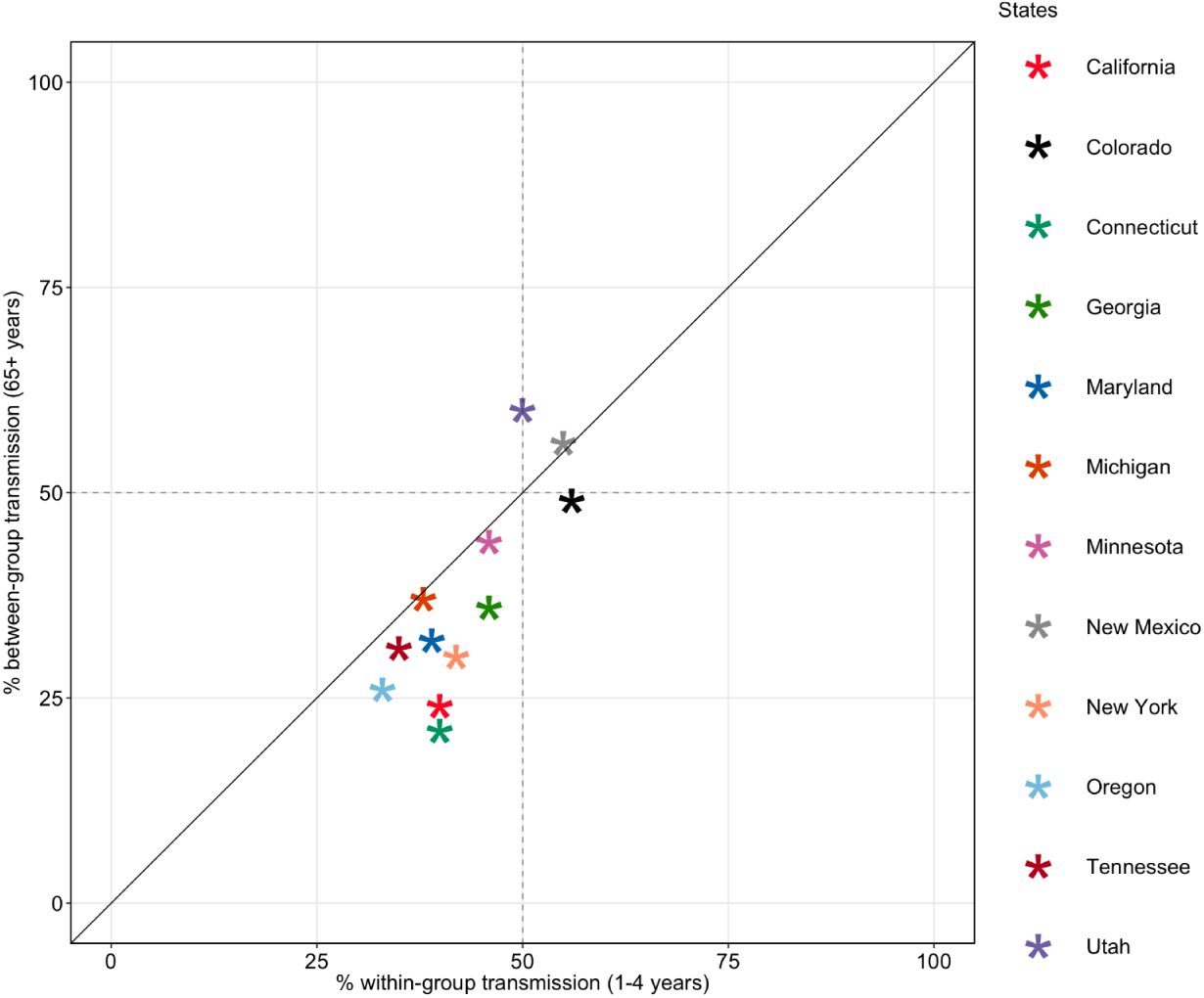
Proportions of within-group and between-group RSV transmission. The estimated hospitalization values from the best-fit model of each state were used to calculate the time-averaged proportions of the mean explained by within-host transmissions in the group of 1-4 years and between-group transmissions in the group of above 65 years. A strong Pearson’s correlation coefficient was found (*r* = 0.81 *p* < 0.005) between the proportion of within-group transmission in the young children group and that of between-group transmission in the elderly group.

**Supplement Table 1.** Estimated parameter values based on the best-fit model to New York data. All parameter values are shown in an exponential form. “*ne.*” indicates the epidemic component for each age group, and “*end*.” indicates the endemic component for each age group. “*overdisp*” is the estimate of the overdispersion parameter. The best-fit model is given by a shared overdispersion parameter among age groups.

| Coefficients | Estimate | Std.Error |
| --- | --- | --- |
| exp(ne.1) | 0.85 | 0.06 |
| exp(ne.1-4) | 0.16 | 0.02 |
| exp(ne.5-17) | 0.02 | 0.00 |
| exp(ne.18-64) | 0.02 | 0.00 |
| exp(ne.above 65) | 0.16 | 0.02 |
| ne.A(2 * pi * t/52).under 1 | 0.32 | 0.07 |
| ne.s(2 * pi * t/52).under 1 | 0.08 | 0.26 |
| ne.A(2 * pi * t/52).1-4 | 0.53 | 0.08 |
| ne.s(2 * pi * t/52).1-4 | 0.22 | 0.17 |
| ne.A(2 * pi * t/52).5-17 | 0.84 | 0.17 |
| ne.s(2 * pi * t/52).5-17 | -0.16 | 0.20 |
| ne.A(2 * pi * t/52).18-64 | 0.36 | 0.12 |
| ne.s(2 * pi * t/52).18-64 | -1.75 | 0.05 |
| ne.A(2 * pi * t/52).above 65 | 0.47 | 0.11 |
| ne.s(2 * pi * t/52).above 65 | -1.64 | 0.05 |
| exp(end.1) | 0.00 | 0.00 |
| exp(end.1-4) | 1.03 | 0.39 |
| exp(end.5-17) | 0.08 | 0.07 |
| exp(end.18-64) | 0.51 | 0.23 |
| exp(end.above 65) | 0.64 | 0.28 |
| exp(end.christmas) | 0.00 | 0.00 |
| exp(end.thanks_giving) | 1.72 | 1.11 |
| exp(end.t) | 1.01 | 0.00 |
| overdisp | 0.15 | 0.02 |

## References

1. Tin Tin Htar M, Yerramalla MS, Moïsi JC, Swerdlow DL. The burden of respiratory syncytial virus in adults: a systematic review and meta-analysis. Epidemiol Infect. 2020; 148:e48.

2. Shi T, McAllister DA, O’Brien KL, et al. Global, regional, and national disease burden estimates of acute lower respiratory infections due to respiratory syncytial virus in young children in 2015: a systematic review and modelling study. Lancet. 2017; 390(10098):946–958.

3. Li Y, Wang X, Blau DM, et al. Global, regional, and national disease burden estimates of acute lower respiratory infections due to respiratory syncytial virus in children younger than 5 years in 2019: a systematic analysis. Lancet. 2022; 399(10340):2047–2064.

4. Zheng Z, Warren JL, Artin I, Pitzer VE, Weinberger DM. Relative timing of respiratory syncytial virus epidemics in summer 2021 across the United States was similar to a typical winter season. Influenza Other Respi Viruses. Wiley; 2022; 16(4):617–620.

5. Pitzer VE, Viboud C, Alonso WJ, et al. Environmental drivers of the spatiotemporal dynamics of respiratory syncytial virus in the United States. PLoS Pathog. 2015; 11(1):e1004591.

6. Goldstein E, Nguyen HH, Liu P, et al. On the Relative Role of Different Age Groups During Epidemics Associated With Respiratory Syncytial Virus. J Infect Dis. academic.oup.com; 2018; 217(2):238–244.

7. Mazur NI, Terstappen J, Baral R, et al. Respiratory syncytial virus prevention within reach: the vaccine and monoclonal antibody landscape. Lancet Infect Dis. Elsevier BV; 2023; 23(1):e2–e21.

8. Löwensteyn YN, Zheng Z, Rave N, et al. Year-Round Respiratory Syncytial Virus Transmission in The Netherlands Following the COVID-19 Pandemic: A Prospective Nationwide Observational and Modeling Study. J Infect Dis. 2023; 228(10):1394–1399.

9. Zheng Z, Pitzer VE, Warren JL, Weinberger DM. Community factors associated with local epidemic timing of respiratory syncytial virus: A spatiotemporal modeling study. Sci Adv. American Association for the Advancement of Science (AAAS); 2021; 7(26):eabd6421.

10. Zheng Z, Weinberger DM, Pitzer VE. Predicted effectiveness of vaccines and extended half-life monoclonal antibodies against RSV hospitalizations in children. NPJ Vaccines. Springer Science and Business Media LLC; 2022; 7(1):127.

11. Meyer S, Held L. Incorporating social contact data in spatio-temporal models for infectious disease spread. Biostatistics. Oxford Academic; 2017; 18(2):338–351.

12. Cori A, Ferguson NM, Fraser C, Cauchemez S. A new framework and software to estimate time-varying reproduction numbers during epidemics. Am J Epidemiol. Oxford University Press (OUP); 2013; 178(9):1505–1512.

13. Vink MA, Bootsma MCJ, Wallinga J. Serial intervals of respiratory infectious diseases: a systematic review and analysis. Am J Epidemiol. Oxford University Press (OUP); 2014; 180(9):865–875.

14. Mistry D, Litvinova M, Pastore Y Piontti A, et al. Inferring high-resolution human mixing patterns for disease modeling. Nat Commun. Springer Science and Business Media LLC; 2021; 12(1):323.

15. Venter M, Madhi SA, Tiemessen CT, Schoub BD. Genetic diversity and molecular epidemiology of respiratory syncytial virus over four consecutive seasons in South Africa: identification of new subgroup A and B genotypes. J Gen Virol. Microbiology Society; 2001; 82(Pt 9):2117–2124.

16. Agoti CN, Munywoki PK, Phan MVT, et al. Transmission patterns and evolution of respiratory syncytial virus in a community outbreak identified by genomic analysis. Virus Evol. Oxford University Press (OUP); 2017; 3(1):vex006.

17. Agoti CN, Phan MVT, Munywoki PK, et al. Genomic analysis of respiratory syncytial virus infections in households and utility in inferring who infects the infant. Sci Rep. Springer Science and Business Media LLC; 2019; 9(1):10076.

18. Langedijk AC, Vrancken B, Lebbink RJ, et al. The genomic evolutionary dynamics and global circulation patterns of respiratory syncytial virus. Nat Commun. Nature Publishing Group; 2024; 15(1):3083.

19. Sun M, Lai H, Na F, et al. Monoclonal antibody for the prevention of respiratory syncytial virus in infants and children: A systematic review and network meta-analysis. JAMA Netw Open. 2023; 6(2):e230023.

20. Langedijk AC, Bont LJ. Respiratory syncytial virus infection and novel interventions. Nat Rev Microbiol. Springer Science and Business Media LLC; 2023; 21(11):734–749.

21. Drysdale SB, Cathie K, Flamein F, et al. Nirsevimab for prevention of hospitalizations due to RSV in infants. N Engl J Med. 2023; 389(26):2425–2435.

22. Kampmann B, Madhi SA, Munjal I, et al. Bivalent prefusion F vaccine in pregnancy to prevent RSV illness in infants. N Engl J Med. 2023; 388(16):1451–1464.

